# Predictive Modeling on the Number of Covid-19 Death Toll in the United States Considering the Effects of Coronavirus-Related Changes and Covid-19 Recovered Cases

**DOI:** 10.1101/2020.06.15.20132357

**Authors:** Hoang Pham

## Abstract

COVID-19 is caused by a coronavirus called SARS-CoV-2. Many countries around the world implemented their own policies and restrictions designed to limit the spread of Covid-19 in recent months. Businesses and schools transitioned into working and learning remotely. In the United States, many states were under strict orders to stay home at least in the month of April. In recent weeks, there are some significant changes related restrictions include social-distancing, reopening states, and staying-at-home orders. The United States surpassed 2 million coronavirus cases on Monday, June 15, 2020 less than five months after the first case was confirmed in the country. The virus has killed at least 115,000 people in the United States as of Monday, June 15, 2020, according to data from Johns Hopkins University.

With the recent easing of coronavirus-related restrictions and changes on business and social activity such as stay-at-home, social distancing since late May 2020 hoping to restore economic and business activities, new Covid-19 outbreaks are on the rise in many states across the country. Some researchers expressed concern that the process of easing restrictions and relaxing stay-at-home orders too soon could quickly surge the number of infected Covid-19 cases as well as the death toll in the United States. Some of these increases, however, could be due to more testing sites in the communities while others may be are the results of easing restrictions due to recent reopening and changed policies, though the number of daily death toll does not appear to be going down in recent days due to Covid-19 in the U.S. This raises the challenging question:

- How can policy decision-makers and community leaders make the decision to implement public policies and restrictions and keep or lift staying-at-home orders of ongoing Covid-19 pandemic for their communities in a scientific way?

In this study, we aim to develop models addressing the effects of recent Covid-19 related changes in the communities such as reopening states, practicing social-distancing, and staying-at-home orders. Our models account for the fact that changes to these policies which can lead to a surge of coronavius cases and deaths, especially in the United States. Specifically, in this paper we develop a novel generalized mathematical model and several explicit models considering the effects of recent reopening states, staying-at-home orders and social-distancing practice of different communities along with a set of selected indicators such as the total number of coronavirus recovered and new cases that can estimate the daily death toll and total number of deaths in the United States related to Covid-19 virus. We compare the modeling results among the developed models based on several existing criteria. The model also can be used to predict the number of death toll in Italy and the United Kingdom (UK). The results show very encouraging predictability for the proposed models in this study.

The model predicts that 128,500 to 140,100 people in the United States will have died of Covid-19 by July 4, 2020. The model also predicts that between 137,900 and 154,000 people will have died of Covid-19 by July 31, and 148,500 to 169,700 will have died by the end of August 2020, as a result of the SARS-CoV-2 coronavirus that causes COVID-19 based on the Covid-19 death data available on June 13, 2020.

The model also predicts that 34,900 to 37,200 people in Italy will have died of Covid-19 by July 4, and 36,900 to 40,400 people will have died by the end of August based on the data available on June 13, 2020. The model also predicts that between 43,500 and 46,700 people in the United Kingdom will have died of Covid-19 by July 4, and 48,700 to 51,900 people will have died by the end of August, as a result of the SARS-CoV-2 coronavirus that causes COVID-19 based on the data available on June 13, 2020.

The model can serve as a framework to help policy makers a scientific approach in quantifying decision-makings related to Covid-19 affairs.

## 1. Introduction

Coronaviruses are a large family of viruses that cause illness ranging from the common cold to more severe diseases. COVID-19, short for “coronavirus disease 2019”, a virus strain, first identified in Wuhan City, Hubei Province, China, that has only spread in people since December 2019. COVID-19 is a new disease and many experts in the field are still learning how it spreads. It has rapidly spread to many countries around the world including the United States [1]. Worldwide travel due to globalization has also results in an increased risk of disease transmission [2]. The outbreak of Covid-19 has rapidly spread to more than 220 countries around the world including the United States.

There were more than 2 million confirmed coronavirus cases on Monday, June 15 and at least 115,000 people have died in the United States according to a tally by Johns Hopkins University [3]. Already in June, about 300,000 new Covid-19 cases and at least 10,000 more have died in the United States as of Monday, June 15, 2020, according to data from Johns Hopkins University. Worldwide, there are about 8 million confirmed cases and more than 430,000 reported deaths as of Monday, June 15, 2020 [3]

Many countries around the world implemented their own policies and restrictions designed to limit the spread of Covid-19 in recent months. In the United States, many states were under strict orders to stay home at least in the month of April. The worst weeks for coronavirus deaths in the U.S. were in early April. In recent weeks, there are some significant developments related restrictions include social-distancing, reopening states, and staying-at-home orders. Some researchers expressed concern the process of easing restrictions and relaxing stay-at-home orders too soon can quickly surge the number of infected Covid-19 cases as well as the death toll in the United States.

COVID-19 is primarily spread through respiratory droplets which means to become infected people generally in within six feet of someone who is contagious and come into contact with these droplets. While the best way to prevent illness is to avoid virus exposure, the Centers for Disease Control and Prevention [4] recommends taking preventive actions to contain the spread of viruses. This includes: (i) avoid touching your eyes, nose, and mouth; (ii) stay home when you are sick; (iii) cover your cough or sneeze with a tissue, then throw the tissue in the trash; (iv) clean and disinfect frequently touched objects and surfaces using a regular household cleaning spray or wipes, and (v) wash your hands often with soap and water for at least 20 seconds, especially after going to the bathroom, before eating, after blowing your nose, coughing, or sneezing. Chin, Chu et al [5] investigated the stability and detection of severe acute respiratory syndrome coronavirus 2 (SARS-CoV 2) in different environmental conditions including temperatures, surfaces. The US Centers for Disease Control and Prevention projected more than 127,000 coronavirus deaths in the country by June 27 [24].

Recently there are some researchers have been developed mathematical models to determine timelines for social distancing, healthcare policies and social distancing practice [6-9], case fatality rate [10-12] and disease transmission [13-14] and disease detection in different environments [5]. Pham [15] recently develops a model that can estimate the cumulative number of deaths in the United States due to the ongoing Covid-19 virus based on a logistic function. This model, however, assumed that there is no significant change in the coming days due to various testing strategies, social-distancing policies, the reopening of community strategies, and staying-at-home orders policy. The model also did not take into account the daily new cases and the total number of new cases of Covid-19. Based on the Covid-19 global and United States death data, it appears that the cumulative number of deaths can follow an S-shaped curves. There are a number of existing S-shaped logistic models in the literature [16-19].

In this paper, we develop a novel generalized mathematical model and several explicit models considering the effects of recent coronavirus-related restrictions include reopening states, staying-at-home orders and social-distancing practice of different communities as well as coronavirus recovered and new cases. The models can estimate the number of daily deaths and total number of death toll in the United States related to Covid-19 virus.

We analyze the proposed models using several existing criteria for the Covid-19 data to predict the death toll in the United States, Italy and UK. The results show that the proposed models fit significantly well. The results show very encouraging predictability for the proposed models in this study. Section 2 discusses the model development and present several explicit models that can estimate the total number of deaths in the US, Italy and UK. Section 3 discusses the modeling results based on the Covid-19 deaths data. Section 4 briefly discuss the findings and conclusion remarks.

## 2. Model Development on Estimating the Number of Deaths

In this section, we discuss the models addressing the effects of recent Covid-19 related changes in the communities such as reopening states, practicing social-distancing, and staying-at-home orders. Our models account for the fact that changes to these policies which can lead to a surge of coronavius cases and deaths, especially in the United States. In other words, we discuss the models with considerations of the effects of recent reopening states, staying-at-home orders and practicing social-distancing of different communities along with a set of selected indicators such as the total number of coronavirus recovered and new cases that can estimate the daily death toll and total number of Covid-19 deaths in the US, Italy and U.K.

### 2.1 Model Considerations

We first present a generalized model addressing the Covid-19 related changes and restrictions such as reopening states, practicing social-distancing, staying-at-home orders along with a set of selected indicators include the total number of cases and the death toll. Specifically, the model takes into considerations that:

1. There are a few people in the communities who have already infected Covid-19 virus from the beginning and are spreading the virus into the community but do not know if they are infected the virus. Consequently, the virus is spreading through the people who are in close contact with one another.
2. The virus is spreading through the areas based on a time-dependent infected rate per person in which it will be spreading at a very slow rate from the beginning due to a small number of infected people and will be spreading at a growth rate much faster due to more number of people who have already infected the virus and are in close contact with non-infected individuals as the time progress, and hopefully with the help of testing strategies and those restrictions designed to limit the spread of Covid-19, then growing slowly until it reaches the maximum daily death toll related to Covid-19 virus.
3. The model also considers the effects of recent Covid-19 related changes such as reopening states, practicing social-distancing, staying-at-home orders along with a set of selected indicators of the Covid-19 data that can lead to a surge of coronavius cases and deaths.
4. We assume deaths are proportional to infections, but with a lag. There can be a significant time lag between when someone is infected and when they die.

### 2.2 Model Development

We now present a novel generalized mathematical model as follows:

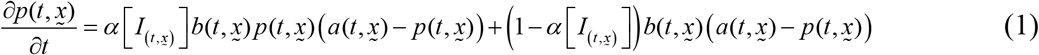

where

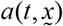 = the total number of deaths plus the death toll to be introduced with respect to 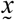 by time t

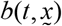 = the time-dependent death rate per person per unit time (i.e., day) w.r.t. 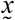

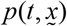 = the cumulative number of deaths by time t w.r.t. 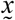

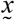 = a set of indicators (i.e., recovered cases, daily new cases, total cases, et al)

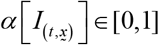 = the overall Covid-19 recovery rate by time t

The general solution 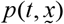 for any given functions 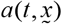 and 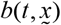 can be found by solving the differential equation from eq. (1). It is expected that the solution for the function 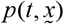 would be very complicated, if the solutions exist. In general, one can develop heuristic algorithms using machine learning approaches that can obtain a heuristic numerical solution for the function 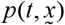 and the model parameter estimates.

Without loss of generality, based on eq. (1) we specifically propose a new simplified model as follows:

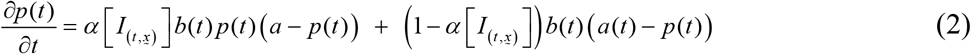

where

*a*(*t*) = the total number of deaths (*a*) plus the death toll due to coronavirus-related restrictions by time t

*b*(*t*) = the time-dependent death rate per person per unit time (i.e., day)

*p*(*t*) = the cumulative number of deaths by time t

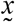 = a set of indicators (i.e., recovered cases, daily new cases, total cases, et al)

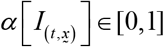 = the overall Covid-19 recovery rate by time t

It is worth noting that the Covid-19 recovery rate 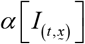 is the function of time t and 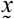, a set of indicators such as recovered cases, daily new Covid-19 cases, total cases etc. We now present some explicit models based on 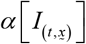 as follows:

*Model 1:* When 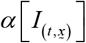 = 1: We propose the following model p(t) that can estimate the cumulative number of deaths at time t:

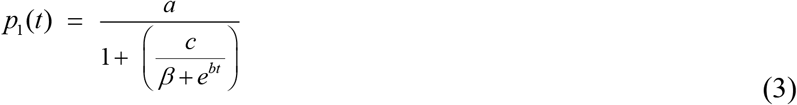

where *a, b, c*, and β are the unknown constant parameters. The daily death toll *r*_1_ (*t*) is given by:

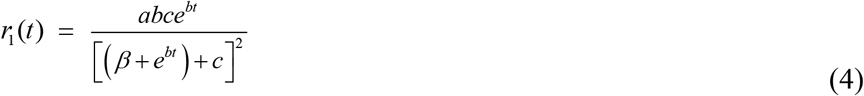

*Model 2:* When 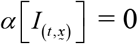: In this case, we present the following model:

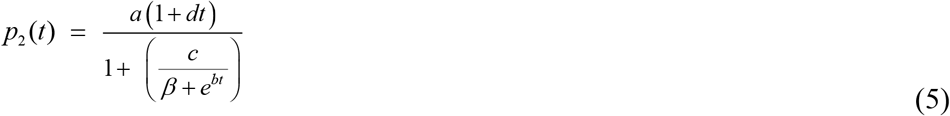

where *a, b, c, d*, and β are the unknown constant parameters The daily death toll *r*_2_ (*t*) can be obtained as follows:

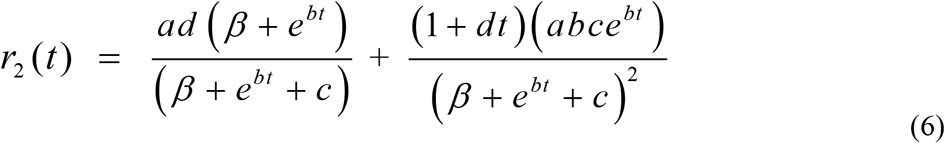

#### 2.2.1 Weighted Recovery Rate Model

When 0 ≤ α ≤ 1: We propose the following weighted recovery rate model:

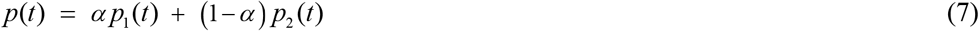

where *p*_*1*_*(t)* and *p*_*2*_*(t)* are given as eq. (3) and (5), respectively. The daily death toll *r*(*t*) can be obtained as follows:

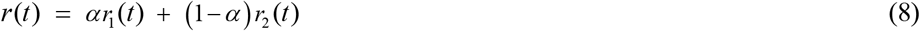

The value 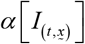 can be estimated based on the number of recovered cases and the total number of Covid-19 cases by the time t.

For various values of 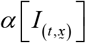, and based on a preliminary study using machine learning approaches, we present the following models in this study:

*Model 3:*

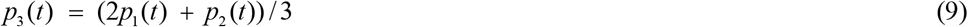

*Model 4:*

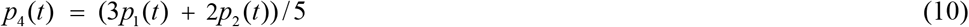

*Model 5:*

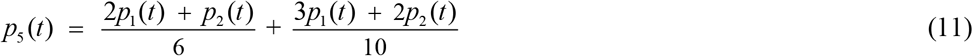

*Model 6:*

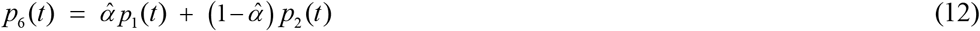

where 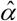 is the empirical estimated parameter based on the recovery rate and the total number of Covid-19 cases.

*Model 7:*

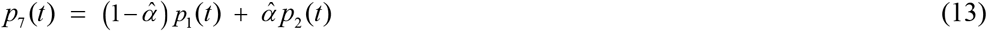

A list of all seven model is shown in Table 1. Here we can estimate these unknown parameters *a, b, c, d* and β using a least squares method and compare their results based on various model criteria such as MSE, AIC, BIC, PC, PIC, PRR and PP (see Table A in the Appendix).

**Table 1.** Models

| Model | $p(t)$ | $r(t)$ |
| --- | --- | --- |
| model 1 | $p_1(t) = \frac{a}{1 + \left( \frac{c}{\beta + e^{bt}} \right)}$ | $r_1(t) = \frac{abce^{bt}}{\left[ (\beta + e^{bt}) + c \right]^2}$ |
| model 2 | $p_2(t) = \frac{a(1+dt)}{1 + \left( \frac{c}{\beta + e^{bt}} \right)}$ | $r_2(t) = \frac{ad(\beta + e^{bt})}{(\beta + e^{bt} + c)} + \frac{(1+dt)(abce^{bt})}{(\beta + e^{bt} + c)^2}$ |
| model 3 | $p_3(t) = (2p_1(t) + p_2(t))/3$ | $r_3(t) = (2r_1(t) + r_2(t))/3$ |
| Model 4 | $p_4(t) = (3p_1(t) + 2p_2(t))/5$ | $r_4(t) = (3r_1(t) + 2r_2(t))/5$ |
| Model 5 | $p_5(t) = \frac{2p_1(t) + p_2(t)}{6} + \frac{3p_1(t) + 2p_2(t)}{10}$ | $r_5(t) = \frac{2r_1(t) + r_2(t)}{6} + \frac{3r_1(t) + 2r_2(t)}{10}$ |
| Model 6 | $p_6(t) = \hat{\alpha} p_1(t) + (1 - \hat{\alpha}) p_2(t)$ | $r_6(t) = \hat{\alpha} r_1(t) + (1 - \hat{\alpha}) r_2(t)$ |
| Model 7 | $p_7(t) = (1 - \hat{\alpha}) p_1(t) + \hat{\alpha} p_2(t)$ | $r_7(t) = (1 - \hat{\alpha}) r_1(t) + \hat{\alpha} r_2(t)$ |

## 3. Modeling Analysis and Prediction Results

In this section, we calculate the total number of deaths and daily death toll for all seven models as shown in Table 1 based on the death data from Worldometer [23] for the United States, Italy and United Kingdom. Worldometer manually analyzes, validates, and aggregates data from thousands of sources in real time and provides global COVID-19 live statistics for people around the world [23]. Worldometer data is also used by many research organizations, government of many countries including the Johns Hopkins CSSE. This Worldometer database provides a comprehensive global data for COVID-19 consisting of many indicators for each countries such as the daily deaths, total deaths, total cases, new cases, active cases, total recovered, serious critical, total cases per one million population, total tests, and total tests per one million population.

### Application 1: COVID-19 Death Data in the United States

We calculate the total number of Covid-19 deaths and daily death toll in the US of all seven models given in Table 1 based on the available data consisting of 106 days obtained from February 29, 2020 to June 13, 2020 as shown in Table 2. Figures 1a and 1b show the real data on the total number of deaths and daily death toll in the US. Table 3 shows the values of model 1 and 2 based on several existing criteria (see Table A in the appendix) such as MSE, AIC, BIC, PC, PIC, PRR and PP. Using Models 6 and 7, we calculate the upper and lower predictions on the number of deaths and the daily death toll in the US.

**Table 2.**
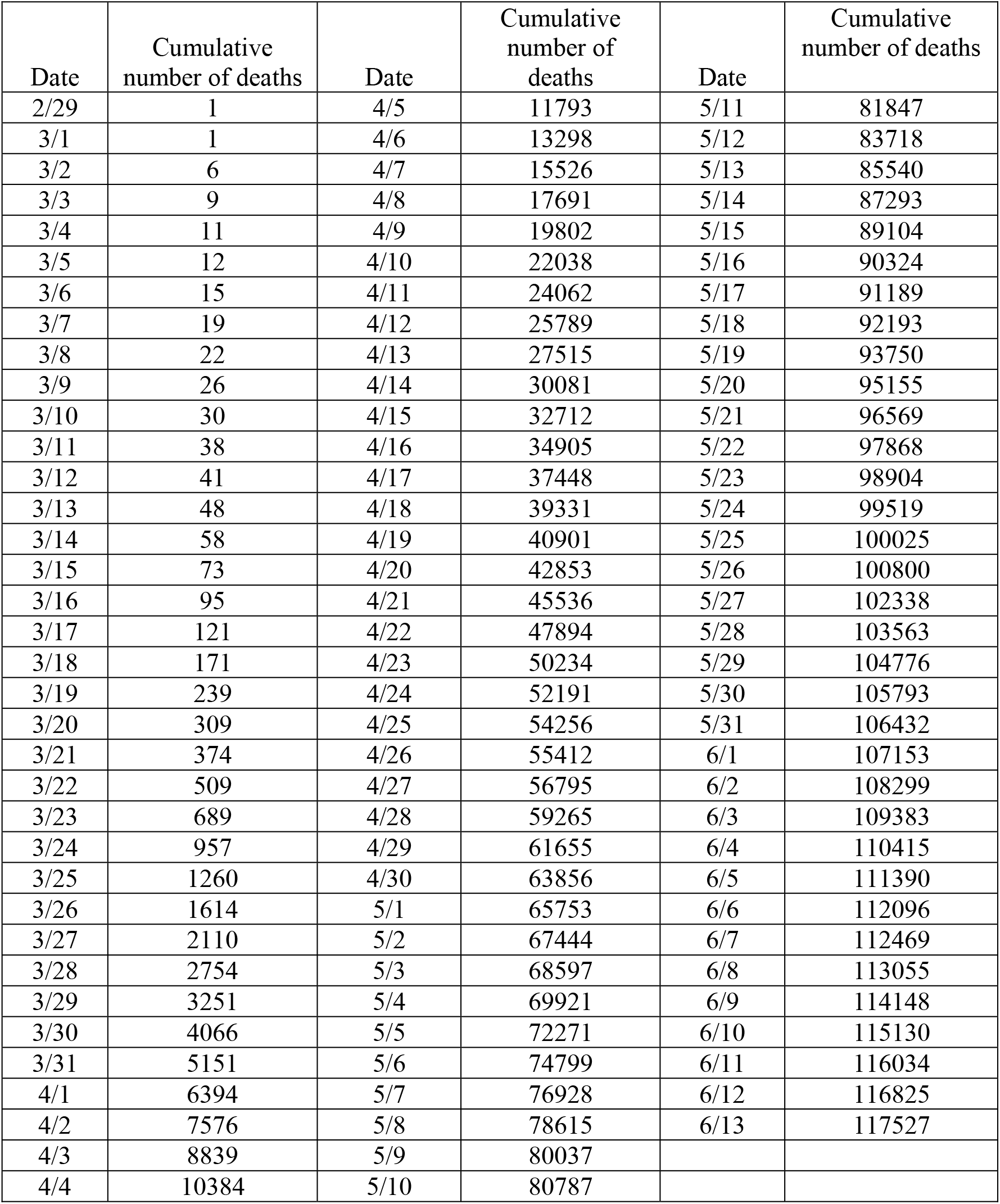
US Deaths data [23] during 2/29/20 - 6/13/20

**Table 3.** Model selection criteria based on available data in the United States

| Selection Criteria | Model 1 | Model 2 |
| --- | --- | --- |
| p(t) | $p_1(t) = \frac{a}{1 + \left( \frac{c}{\beta + e^{bt}} \right)}$ | $p_2(t) = \frac{a(1+dt)}{1 + \left( \frac{c}{\beta + e^{bt}} \right)}$ |
| MSE | 119671330 | 2389277 |
| AIC | 1975.6 | 1561.65 |
| BIC | 1986.2 | 1574.97 |
| PC | 950.77 | 744.43 |
| PIC | 12206475707 | 241316954 |
| PRR | 36.2 | 24.78 |
| PP | 162855.7 | 188284.4 |

**Figure 1a:**
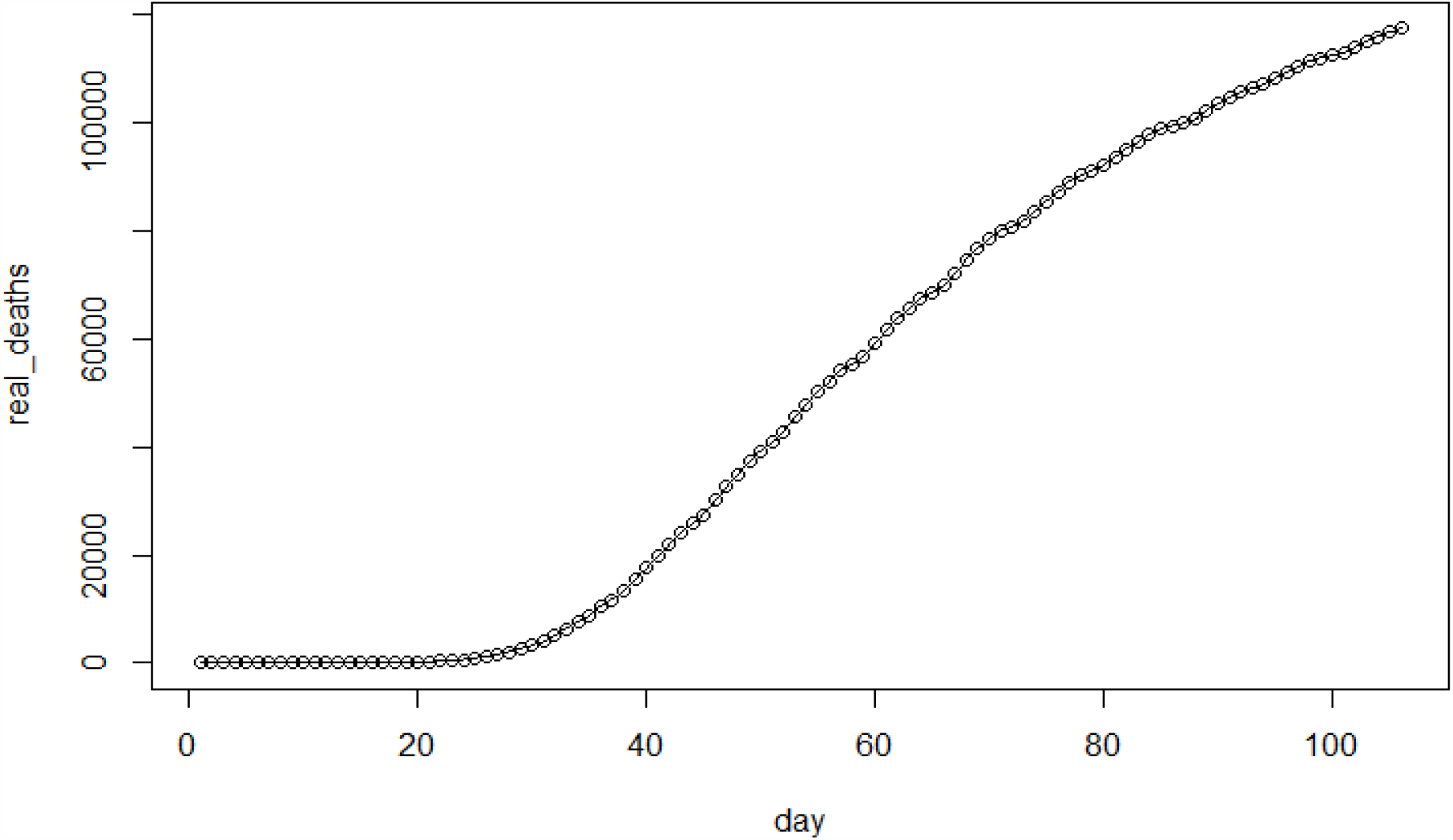
Total death toll vs.day in the United States

**Figure 1b:**
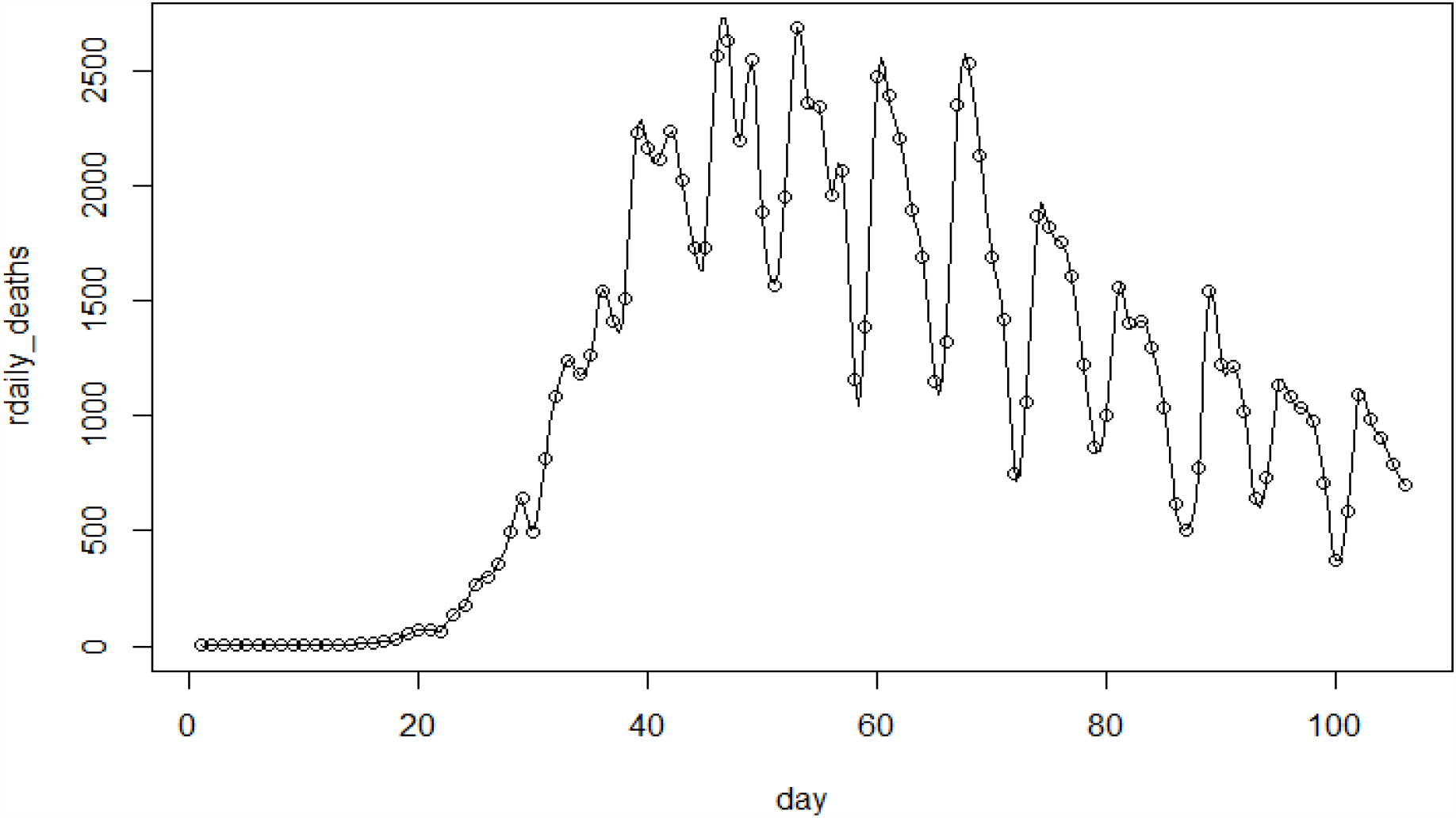
Daily death toll vs.day in the United States

The model predicts that 128,500 to 140,100 people in the United States will have died of Covid-19 by July 4. The model also predicts that between 137,900 and 154,000 people will have died of Covid-19 by July 31, and 148,500 to 169,700 will have died by the end of August 2020, as a result of the SARS-CoV-2 coronavirus that causes COVID-19 based on the Covid-19 death data available on June 13, 2020. Figures 2 and 3 show the prediction on the total number of people and the number of people daily who have died of Covid-19 in the US, respectively. The results show very encouraging predictability for the proposed models. The model can serve as a framework to help policy makers a scientific approach in quantifying decision-makings related to Covid-19 affairs.

**Figure 2:**
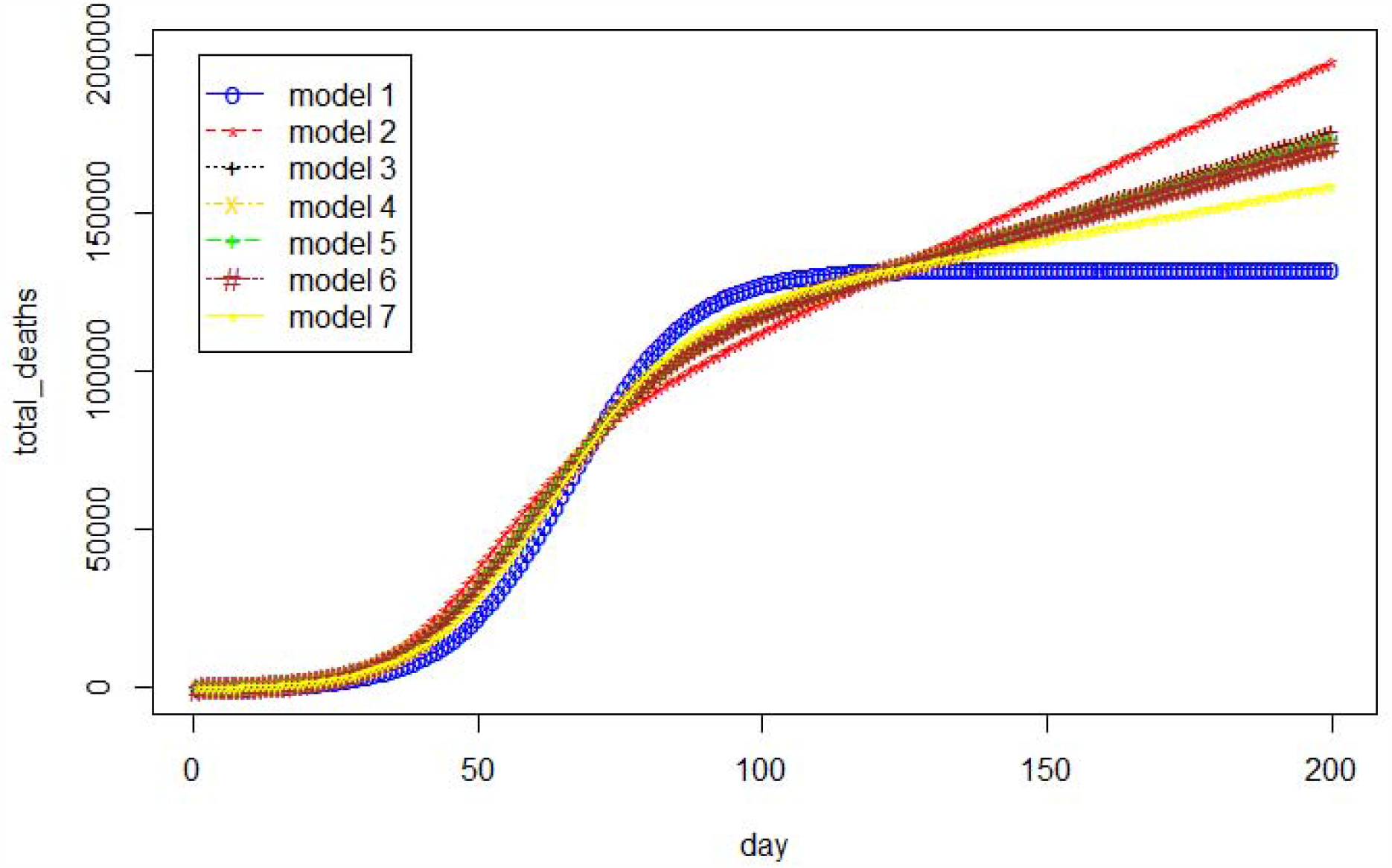
The estimated cumulative number of deaths vs. day in the United States

**Figure 3:**
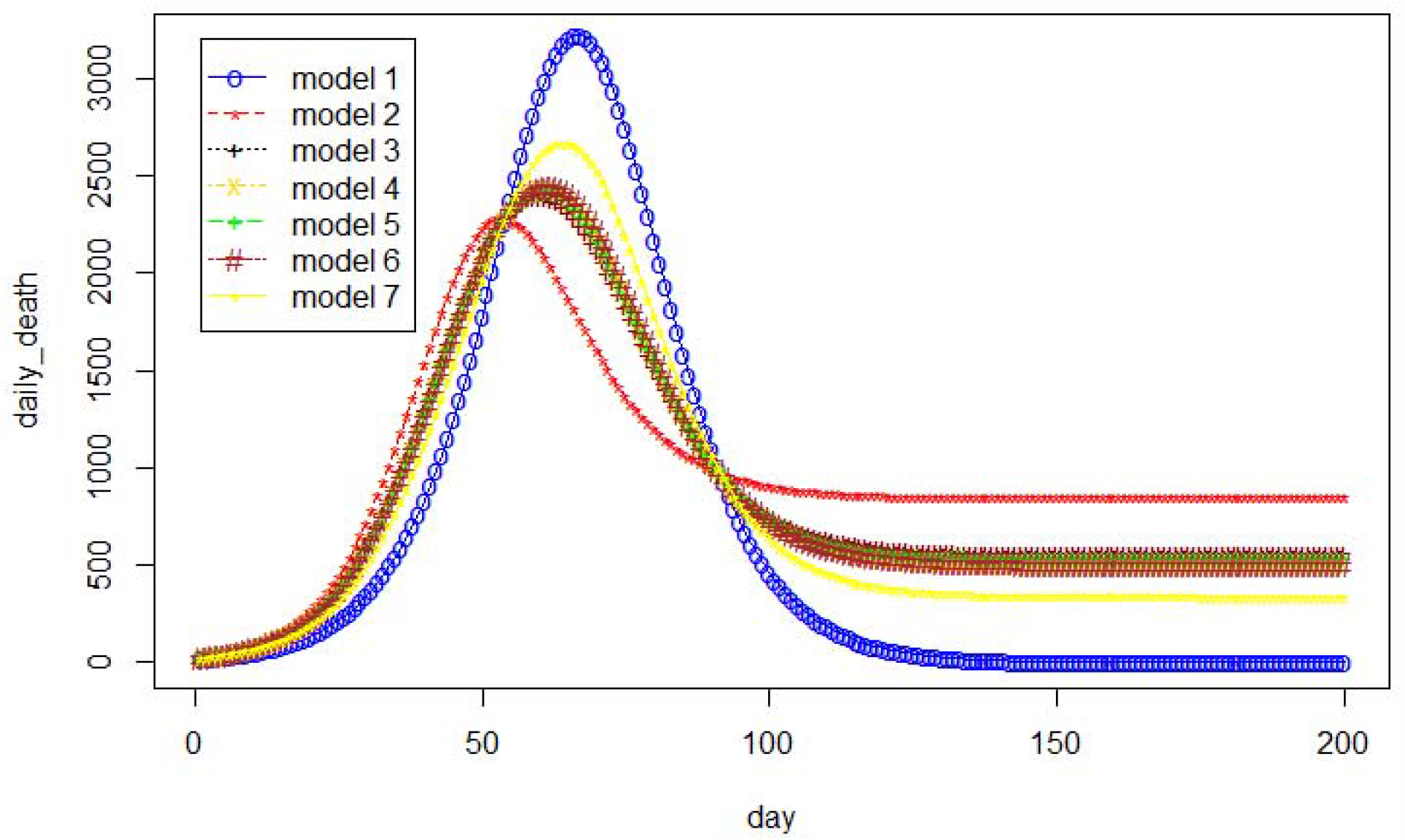
The estimated daily number of deaths vs. day in the United States

### Application 2: COVID-19 Death Data in Italy

We calculate the total number of Covid-19 deaths and daily death toll in Italy of all seven models given in Table 1 based on the available data consisting of 114 days from February 21, 2020 to June 13, 2020 [23]. Similarly, Figures 4a and 4b show the real total number of deaths and daily death toll data in Italy. Table 4 shows the values of model 1 and 2 based on several existing criteria (see Table A in the appendix). From Model 6, we can calculate the upper and lower predictions on the number of deaths and the daily death toll.

**Table 4.**
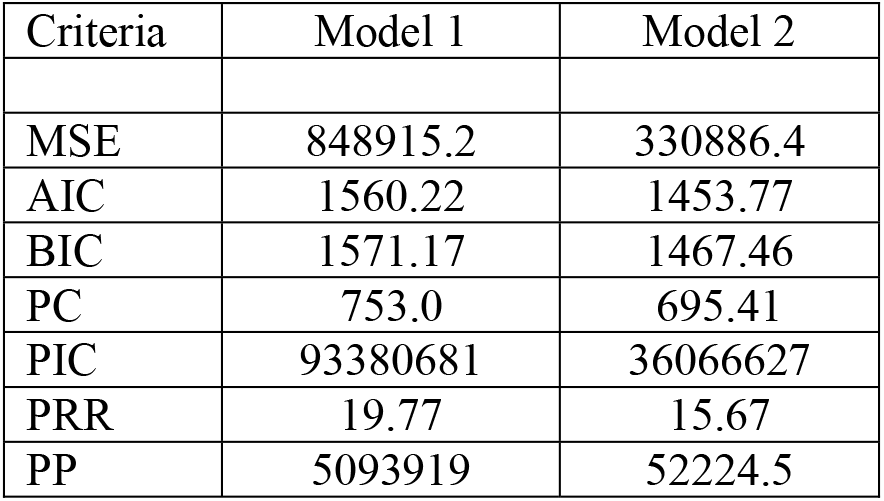
Model selection criteria based on available data in Italy

| Criteria | Model 1 | Model 2 |
| --- | --- | --- |
| MSE | 848915.2 | 330886.4 |
| AIC | 1560.22 | 1453.77 |
| BIC | 1571.17 | 1467.46 |
| PC | 753.0 | 695.41 |
| PIC | 93380681 | 36066627 |
| PRR | 19.77 | 15.67 |
| PP | 5093919 | 52224.5 |

**Figure 4a:**
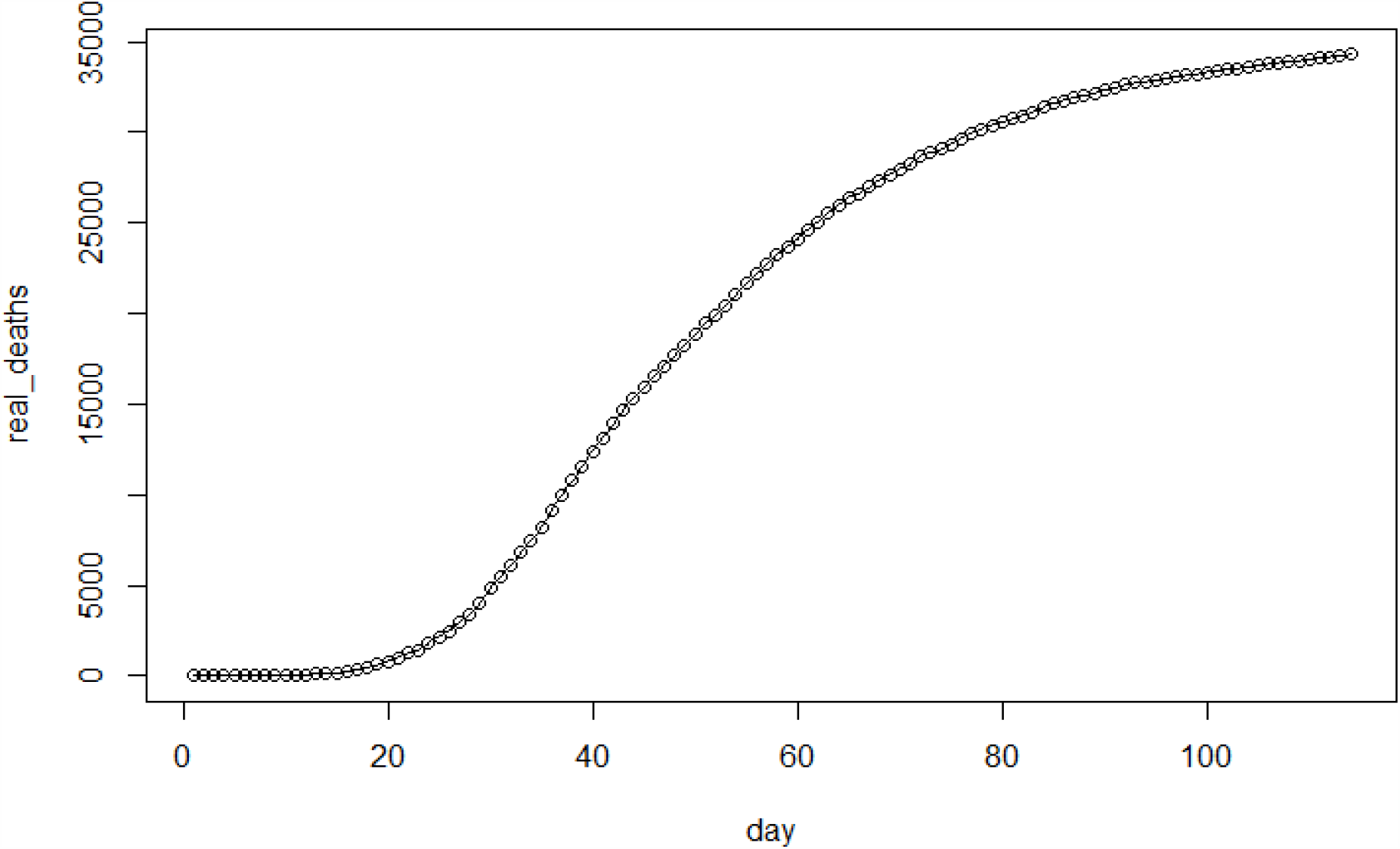
Total death toll vs.day in Italy

**Figure 4b:**
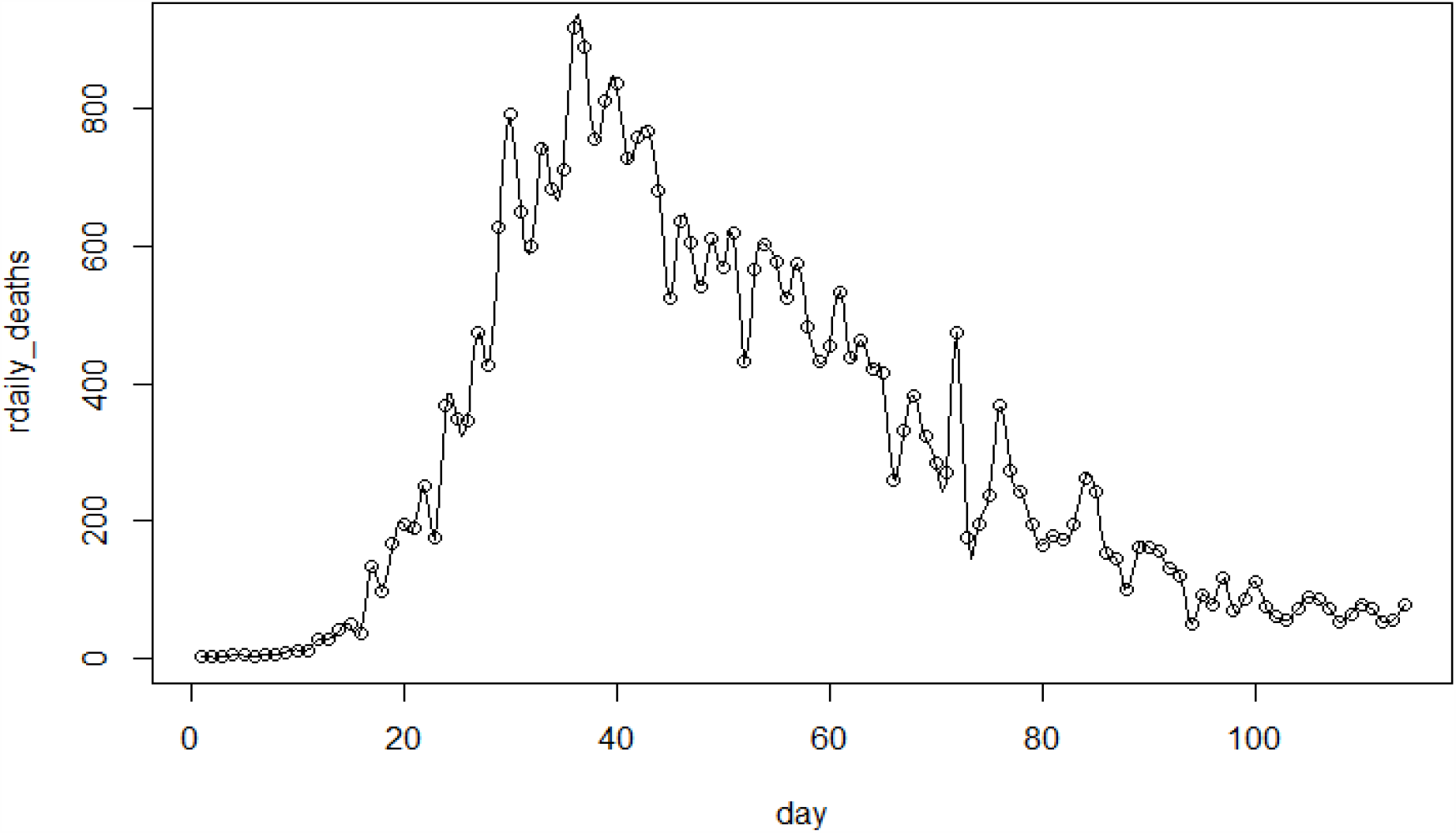
Daily death toll vs.day in Italy

The model predicts that 34,900 to 37,200 people in Italy will have died of Covid-19 by July 4, and 36,900 to 40,400 people will have died by the end of August 2020 based on the data available on June 13, 2020, as a result of the SARS-CoV-2 coronavirus that causes COVID-19 based on the data available on June 13. Figures 5 and 6 show the predictions on the number of people and the number of people daily who have died of Covid-19 in Italy, respectively. The results show very encouraging predictability for the proposed models.

**Figure 5:**
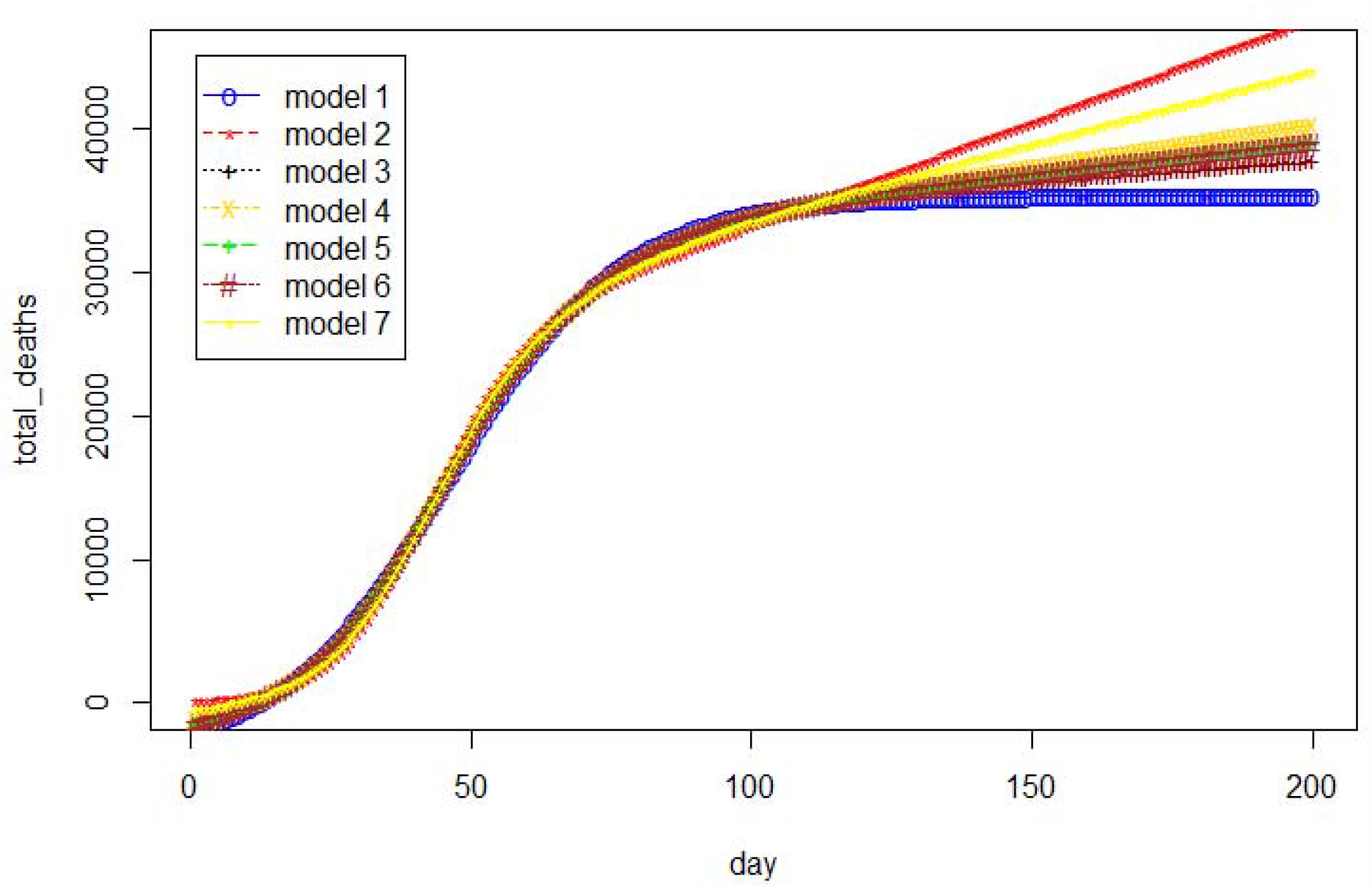
The estimated cumulative number of deaths vs.day in Italy

**Figure 6:**
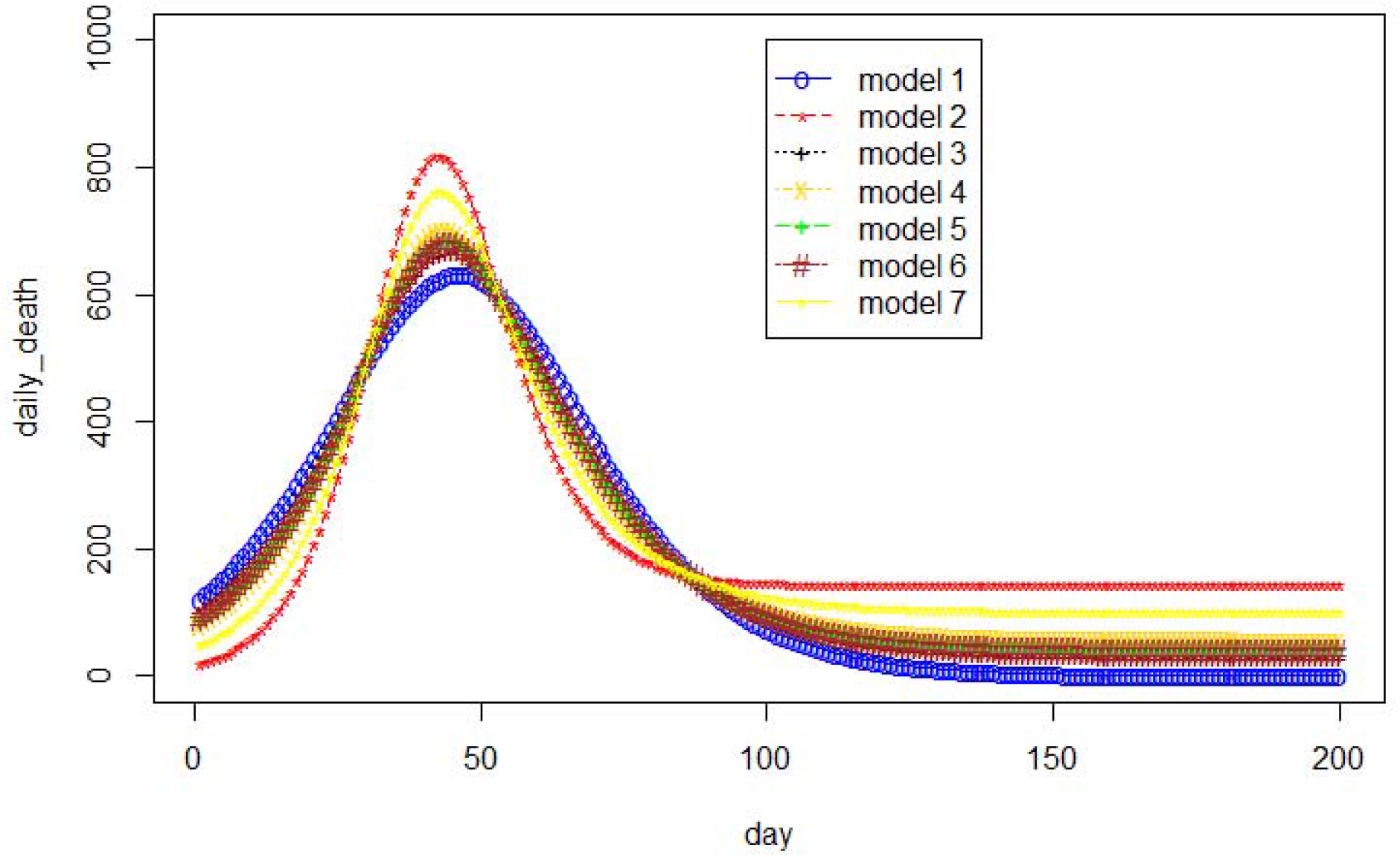
The estimated daily number of deaths vs.day in Italy

### Application 3: COVID-19 Death Data in the United Kingdom

We now calculate the total number of Covid-19 deaths and daily death toll in the United Kingdom of all seven models given in Table 1 based on the available data consisting of 101 days from March 5, 2020 to June 13, 2020 [23]. Figures 7a and 7b show the real total number of deaths and daily death toll data in the U.K. Table 5 shows the values of model 1 and 2 based on several existing criteria. From Model 6, we can obtain the upper and lower predictions on the number of deaths and the daily death toll in the U.K.

**Table 5.**
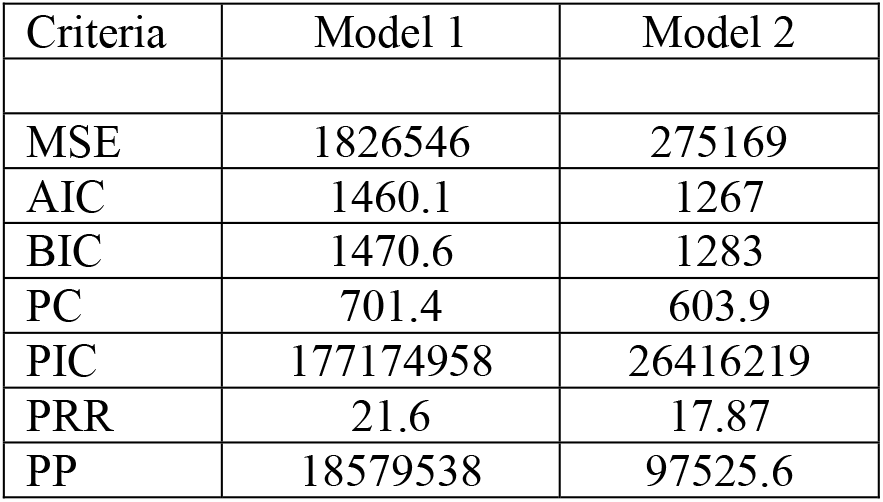
Model selection criteria based on available data in the United Kingdom

**Figure 7a:**
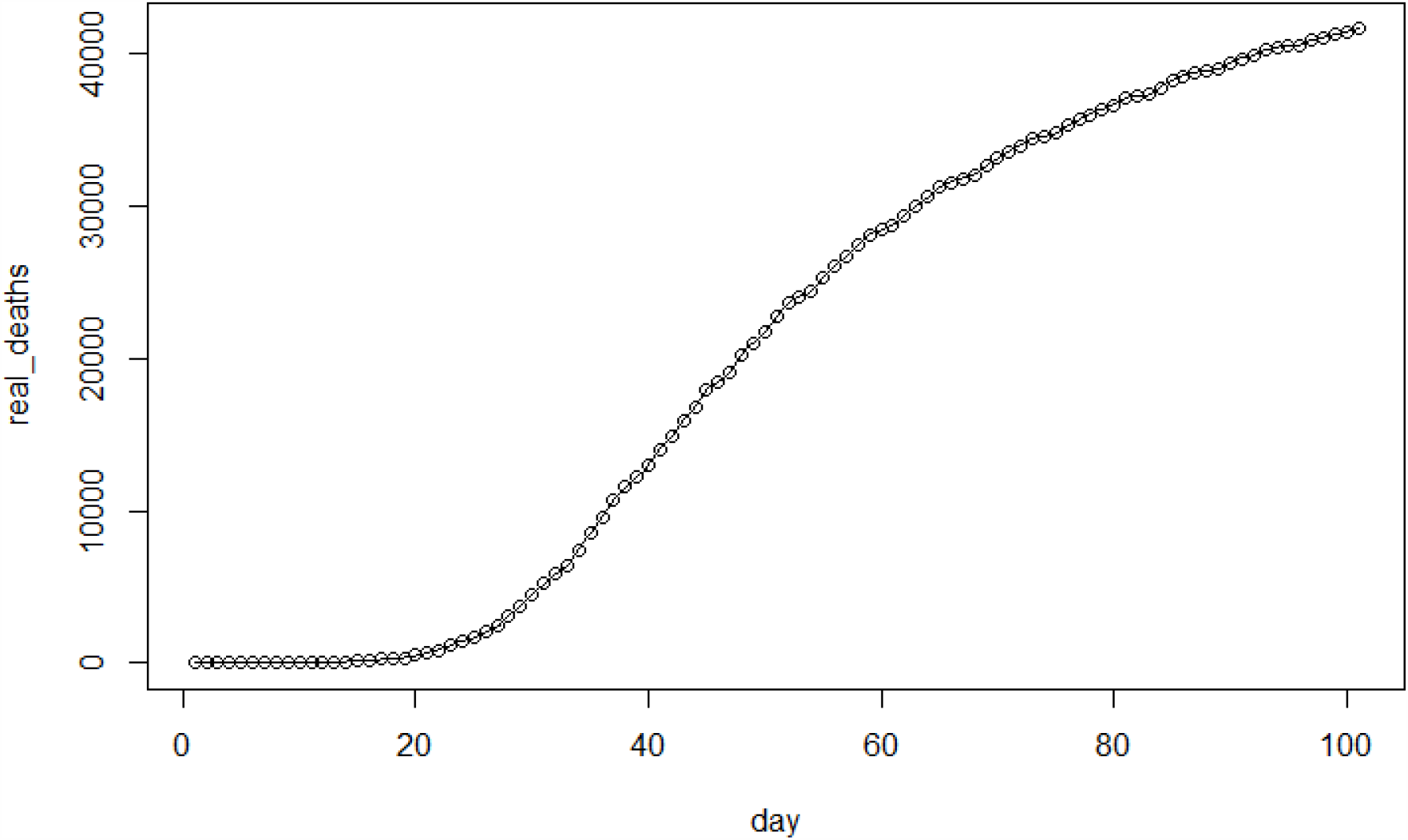
Total death toll vs.day in United Kingdom

**Figure 7b:**
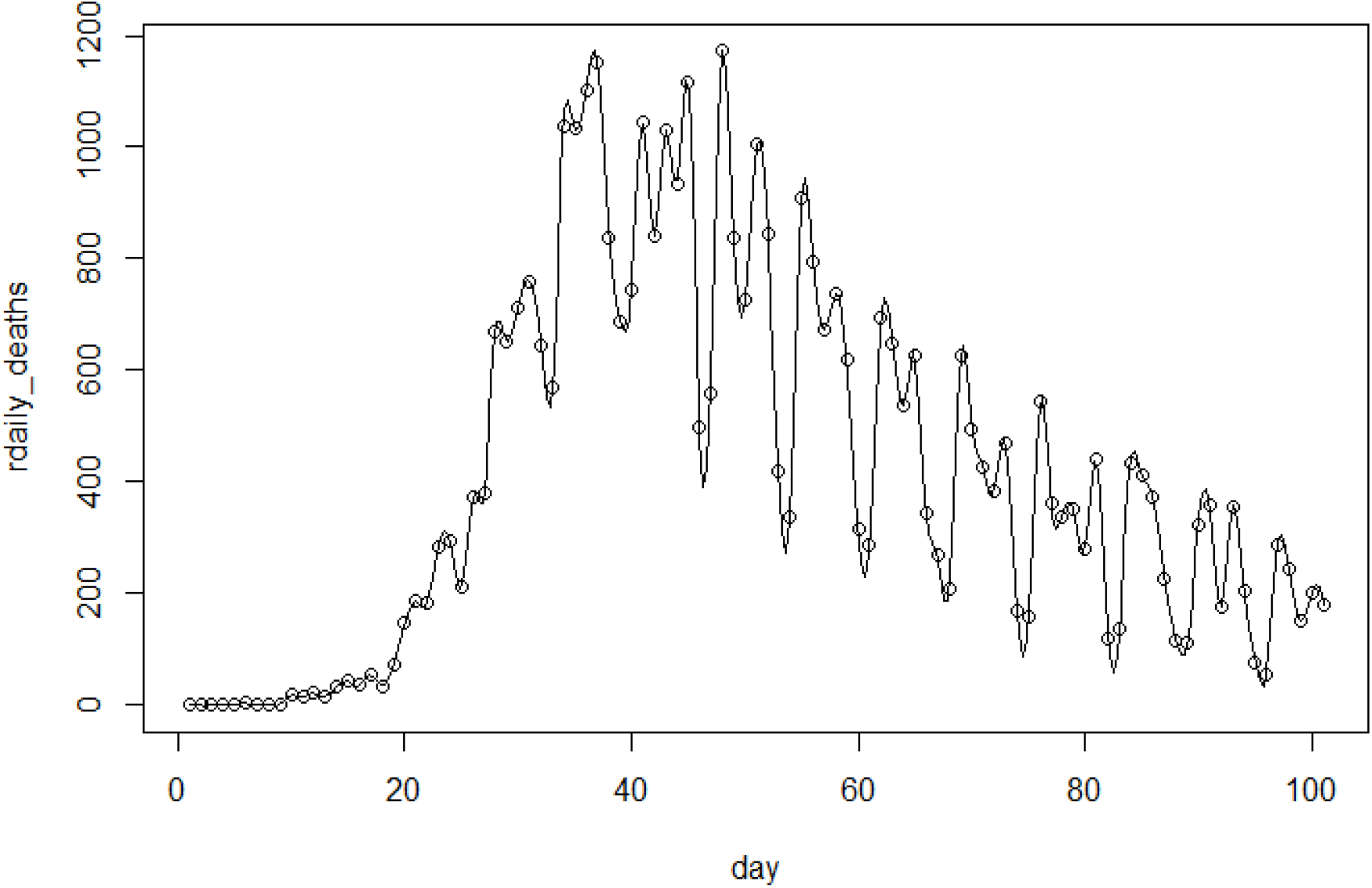
Daily death toll vs.day in United Kingdom

The model also predicts that between 43,500 and 46,700 people in the United Kingdom will have died of Covid-19 by July 4, and 48,700 to 51,900 people will have died by the end of August 2020, as a result of the SARS-CoV-2 coronavirus that causes COVID-19 based on the data available on June 13, 2020. Figures 8 and 9 show the predictions on the number of people and the number of people daily who have died of Covid-19 in the U.K., respectively. The results show very encouraging predictability for the proposed models.

**Figure 8:**
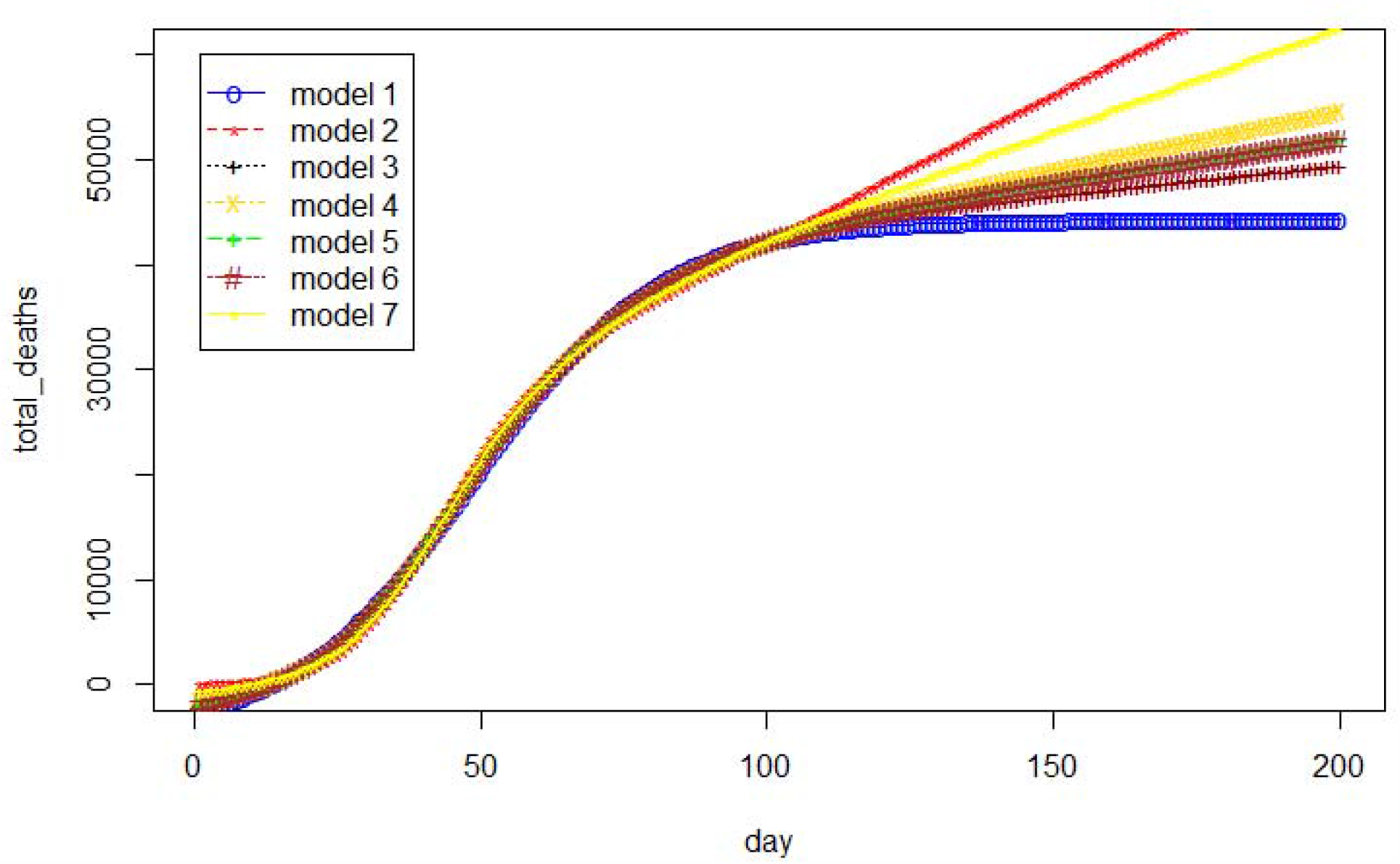
The estimated cumulative number of deaths vs.day in the United Kingdom (UK)

**Figure 9:**
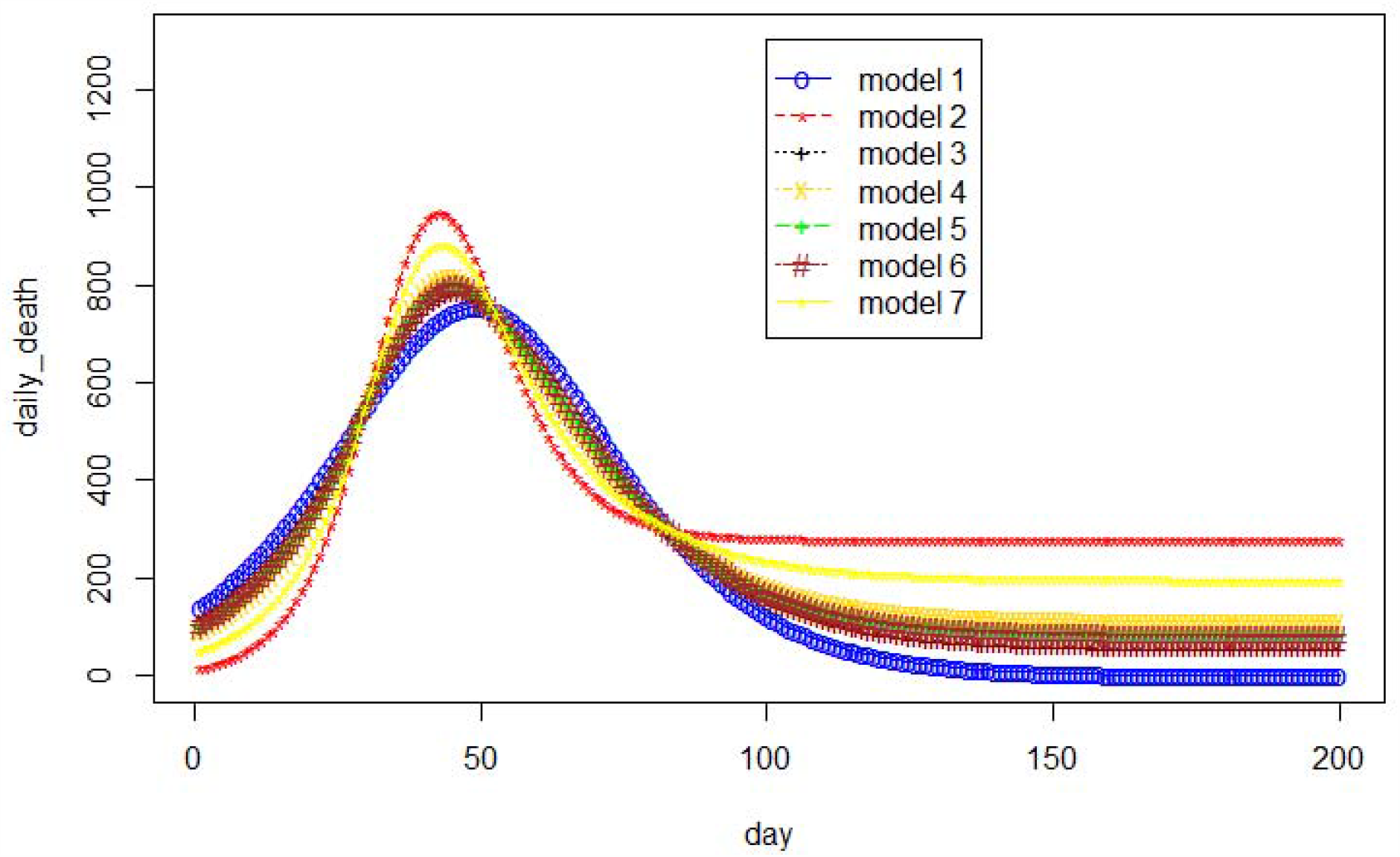
The estimated daily number of deaths vs.day in the United Kingdom (UK)

## 4. Conclusion

This paper presents a generalized mathematical model and several explicit models considering the effects of coronavirus-related restrictions and changes such as reopening states, staying-at-home orders and social-distancing practice of different communities along with a set of selected indicators such as the total number of coronavirus recovered cases and new cases in the US and other countries such as Italy and the United Kingdom. The models can predict the daily death toll and total number of deaths in the US related to Covid-19 virus. The models also predict the daily death toll and total number of deaths in Italy and the United Kingdom. The results show very encouraging predictability for the proposed models. The models can serve as a framework to help policy makers a scientific approach in quantifying decision-makings related to Covid-19 affairs.

## Data Availability

see the link below

https://www.worldometers.info/coronavirus/

## APPENDIX

**Table A.**
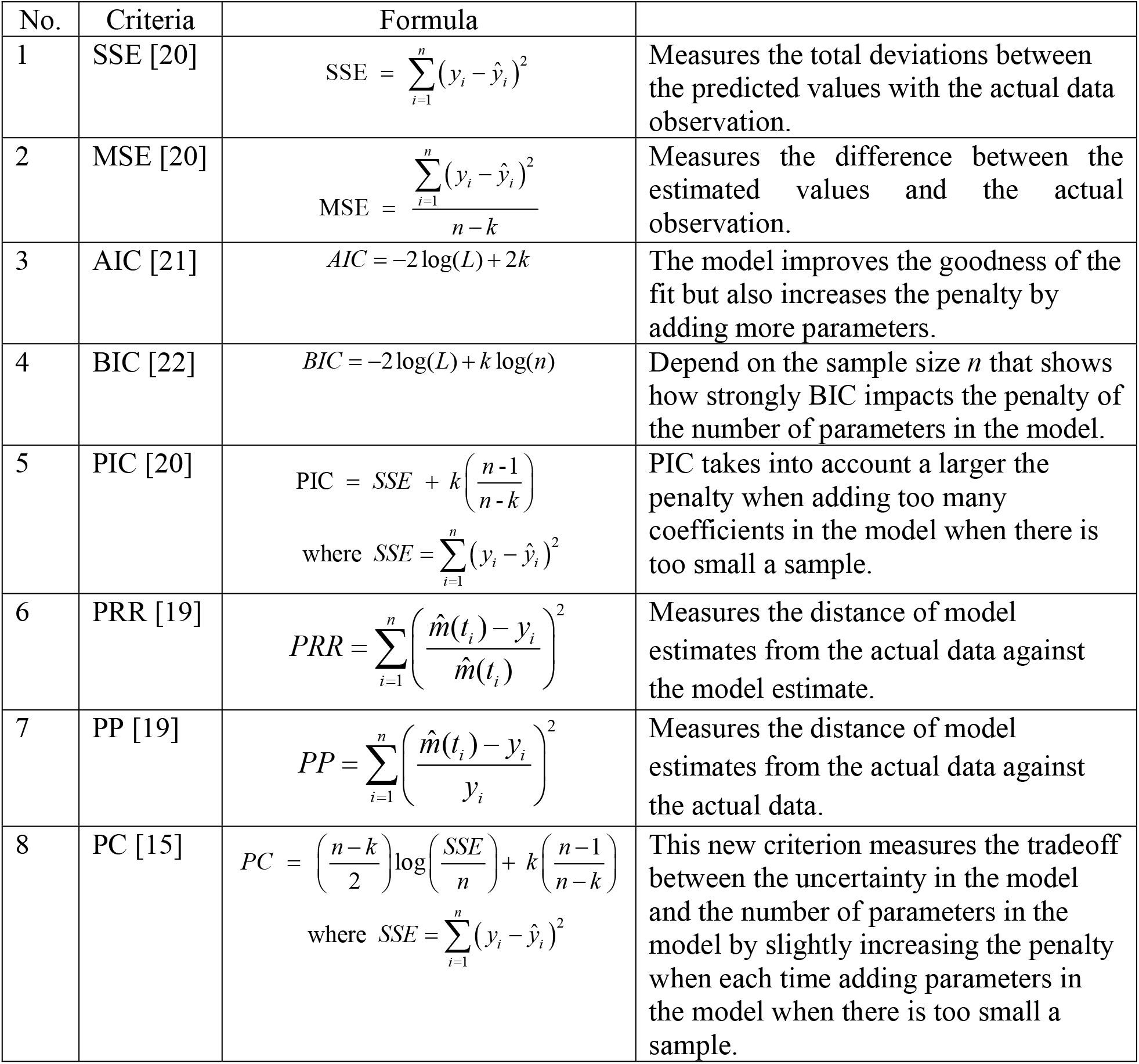
Some Criteria Model Selection

| No. | Criteria | Formula |  |
| --- | --- | --- | --- |
| 1 | SSE [20] | $SSE = \sum_{i=1}^n (y_i - \hat{y}_i)^2$ | Measures the total deviations between the predicted values with the actual data observation. |
| 2 | MSE [20] | $MSE = \frac{\sum_{i=1}^n (y_i - \hat{y}_i)^2}{n - k}$ | Measures the difference between the estimated values and the actual observation. |
| 3 | AIC [21] | $AIC = -2 \log(L) + 2k$ | The model improves the goodness of the fit but also increases the penalty by adding more parameters. |
| 4 | BIC [22] | $BIC = -2 \log(L) + k \log(n)$ | Depend on the sample size $n$ that shows how strongly BIC impacts the penalty of the number of parameters in the model. |
| 5 | PIC [20] | $PIC = SSE + k \left( \frac{n-1}{n-k} \right)$<br>where $SSE = \sum_{i=1}^n (y_i - \hat{y}_i)^2$ | PIC takes into account a larger the penalty when adding too many coefficients in the model when there is too small a sample. |
| 6 | PRR [19] | $PRR = \sum_{i=1}^n \left( \frac{\hat{m}(t_i) - y_i}{\hat{m}(t_i)} \right)^2$ | Measures the distance of model estimates from the actual data against the model estimate. |
| 7 | PP [19] | $PP = \sum_{i=1}^n \left( \frac{\hat{m}(t_i) - y_i}{y_i} \right)^2$ | Measures the distance of model estimates from the actual data against the actual data. |
| 8 | PC [15] | $PC = \left( \frac{n-k}{2} \right) \log \left( \frac{SSE}{n} \right) + k \left( \frac{n-1}{n-k} \right)$<br>where $SSE = \sum_{i=1}^n (y_i - \hat{y}_i)^2$ | This new criterion measures the tradeoff between the uncertainty in the model and the number of parameters in the model by slightly increasing the penalty when each time adding parameters in the model when there is too small a sample. |

## Abbreviations

SSE: sum of squared error
MSE: mean squared error
AIC: Akaike’s information criterion
BIC: Bayesian information criterion
LSE: least squared estimate
PC: Pham’s criterion
PP: the predictive power
PIC: Pham’s information criterion
PRR: the predictive ratio-risk

